# Risk factors among Black and White COVID-19 patients from a Louisiana Hospital System, March, 2020 – August, 2021

**DOI:** 10.1101/2022.07.27.22278118

**Authors:** Qingzhao Yu, Wentao Cao, Diana Hamer, Norman Urbanek, Susanne Straif-Bourgeois, Stephania Cormier, Tekeda Ferguson, Jennifer Richmond-Bryant

**Affiliations:** Biostatistics Program, Louisiana State University Health Sciences Center, New Orleans, LA 70112; Division of Academic Affairs, Our Lady of the Lake Regional Medical Center, Baton Rouge, LA 70808; Department of Forestry and Environmental Resources, North Carolina State University, Raleigh, NC 27695; Epidemiology Program, Louisiana State University Health Sciences Center, New Orleans, LA 70112; Department of Biological Sciences, Louisiana State University, Baton Rouge, LA 70803; Pennington Biomedical Research Center, Baton Rouge, LA 70808

## Abstract

**Objectives:** To investigate relationships between race and COVID-19 hospitalizations, intensive care unit (ICU) admissions, and mortality over time and which characteristics, may mediate COVID-19 associations.

**Methods:** We analyzed hospital admissions, ICU admissions, and mortality among positive COVID-19 cases within the ten-hospital Franciscan Ministries of Our Lady Health System around the Mississippi River Industrial Corridor in Louisiana over four waves of the pandemic from March 1, 2020 – August 31, 2021. Associations between race and each outcome were tested, and multiple mediation analysis was performed to test if other demographic, socioeconomic, or air pollution variables mediate the race-outcome relationships.

**Results:** Race was associated with each outcome over the study duration and during most waves. Early in the pandemic, hospitalization, ICU admission, and mortality rates were greater among Black patients, but as the pandemic progressed these rates became greater in White patients. However, Black patients were still disproportionately represented in these measures. Age was a significant mediator for all outcomes across waves, while comorbidity and emissions of naphthalene and chloroprene acted as mediators for the full study period.

**Conclusions:** The role of race evolved throughout the pandemic in Louisiana, but Black patients bore a disproportionate impact. Naphthalene and chloroprene air pollution partially explained the long-term associations. Our findings imply that air pollution might contribute to the increased COVID-19 hospitalizations and mortality among Black residents in Louisiana but likely do not explain most of the effect of race.

**What is already known on this topic:** Early in the pandemic, there was evidence of disparities in COVID-19 cases, hospitalizations, intensive care unit (ICU) admissions, and mortality due to race. Studies were emerging to indicate that strength of these relationships was waning over time.

**What this study adds:** This study tests relationships between race and hospitalizations, ICU admissions, and mortality and finds that Black patients continue to be disproportionately represented, although that inequity diminished over time. This study, the first to use multiple mediation analysis to study COVID-19 associations, suggests that the relationship between race and health outcome can be explained by mediators including age and, to a lesser extent, comorbidity and air pollution.

**How this study might affect research, practice or policy:** This study supports the need for healthcare resources to be available to Louisiana’s communities of color, for policy to support increased access to health care in the Industrial Corridor region, and for policy to support the reduction of air pollution emissions to disproportionately impacting the health of the Industrial Corridor’s communities of color.

## Introduction

Coronavirus Disease 2019 (COVID-19) severity and mortality have been associated with several vulnerability factors, including comorbidities, environmental exposures, natural disasters, sociodemographic factors, and residence in congregate settings[1,2]. During the first wave of COVID-19 cases in the U.S., transmission among residents of congregate settings was responsible for disease spread[3], while comorbidities among older residents likely elevated risk of death[4]. The second wave of COVID-19 cases in the U.S. saw disproportionate numbers of severe disease and deaths among Black, LatinX, Native American, and immigrant population groups[2,5,6]. The third wave may have occurred in part due to asymptomatic transmission in congregate settings including prisons and long-term care facilities, disproportionately impacting Black and LatinX populations[2].

Soon after the start of the pandemic, some evidence emerged of an association between long-term average air pollution concentrations and prevalence or severity of COVID-19. Notably, significant associations were observed for long-term average particulate matter (PM) having diameter smaller than 2.5 μm (PM_2.5_) concentration with COVID-19 infection[7-9], COVID-19 prevalence[10], intensive care unit (ICU) admission[11,12], ventilator use[12], and mortality[7,11-13]. Associations were also observed for long-term average diesel PM concentration estimates for COVID-19 prevalence and mortality[7]; average nitrogen dioxide (NO_2_) concentrations for prevalence[9,10,14], hospitalization[12], ICU admission[12], ventilator use[12], and mortality[12,14]; ozone (O_3_) concentration for mortality[12]; and hazardous air pollutant indices for respiratory and immunological hazard and mortality[15]. Chen et al.[12] also calculated associations with hospitalization, ICU admission, ventilator use, and mortality for 1-month average concentrations of PM_2.5_ and NO_2_. However, evidence was mixed, with some studies showing no association for NO_2_[11]; O_3_[7,9,10,14]; or PM_2.5_[14,15].

Although many studies suggested a relationship between air pollutant concentration and COVID-19 outcomes, these studies primarily occurred early in the pandemic. Less is known about the association between air pollutant exposure and COVID-19 over time. Sidell et al.[9] studied how the relationship between air pollution and COVID-19 infection changed in a Southern California cohort over four waves spanning March 1, 2020 through February 28, 2021. They observed associations to persist for each wave and the entire duration of their study for both 1-month average and 1-year average PM_2.5_ and NO_2_ concentrations and between COVID-19 infection and 1-year average O_3_ concentrations for the second, third, and fourth waves and entire study duration. However, the magnitude of the associations declined over the third and fourth waves, especially for PM_2.5_. Uncertainties persist about the influence of air pollution on COVID-19 outcomes over the course of the pandemic.

Strategies to respond effectively to public health emergencies such as the COVID-19 pandemic require understanding potential causal pathways for disease outcomes[16,17]. Mediation models can be useful to test how conditions present in populations may influence disease status. For a proposed causal pathway, a directed acyclic graph (DAG) may be used to represent the proposed direct exposure-response pathway and mediating factors that comprise the total effect. For each pathway, linear regression models would then be assigned to represent each pathway in the DAG, and total effect includes the effect of each mediator and direct effect of the predictor. Disparities in COVID-19 outcomes by race combined with evidence about the relationship between COVID-19 and comorbidities, insurance status, and pollution exposure led to the hypothesis that there is a causal pathway between race and COVID-19 mediated by comorbidities, insurance status, and pollution exposure (Supplemental Figure A).

Louisiana parishes routinely score well below the national average for quality of life, morbidity, and mortality indices such as low birthweight, child poverty, and median household income[18]. Based on most recently available data, Louisiana ranks 46^th^ among the states in air quality given by average daily PM_2.5_, 47^th^ in percent smokers among adults, and 45^th^ in COVID-19 death rate. For the period of March 1, 2020 – August 31, 2021, 37.7% of Louisiana’s COVID-19 deaths occurred in people identifying as non-Hispanic Black (hereafter referred to as “Black patients”)[19]; in 2020, that proportion was 41.7%, compared to 31.2% of Louisiana residents identifying as Black[20]. A recent analysis connected disparities, systemic racism, economic stress, and COVID-19 mortality[21].

Given the disproportionate impact of COVID-19 on communities of color in Louisiana and the U.S., the goals of this research are to investigate the relationships of race and COVID-19 outcomes over time and to identify which characteristics, if any, may mediate associations of race with COVID-19 hospitalizations, ICU admissions, and mortality, using individual-level data from a Louisiana hospital system. We investigate factors including race, insurance status, comorbidity, and pollutant exposure for four waves of COVID-19 between March 1, 2020 and August 31, 2021. Identification of differences among these patient cohorts will enable improved approaches to prevention and treatment of COVID-19 for each cohort and may identify structural vulnerabilities for future pandemics.

## Methods

We used the Franciscan Missionaries of Our Lady Health System (FMOLHS) COVID-19 registry to identify patients at ten locations. 13,454 patients ages eighteen years or older who tested positive by a polymerase chain reaction (PCR) COVID-19 test were identified using the Epic healthcare software between March 1, 2020 and August 31, 2021. This period is broken down by waves: March 1 – June 10, 2020, June 11 – October 6, 2020, October 7, 2020 – June 30, 2021, and July 1 – August 31, 2021. These waves were chosen to minimize both cases and mortality at the beginning and end of each period using the Johns Hopkins database for Louisiana[22]. Patient-level variables included hospital department, COVID-19 test date, COVID-19 test result, age, insurance status (private insurance, Medicaid, Medicare, and self-pay), race, ethnicity, sex, admission date, discharge date, length of hospital stay, admission status, ICU stay, ICU admission date, ICU discharge date, length of ICU stay, discharge dispatch, body mass index (BMI), comorbidities, census tract, and census block group. To minimize bias in the patient database, negative PCR tests were not included in the database because tests were often obtained for non-medical reasons (e.g., work, travel, recreation).

Air pollution burden calculations were based on Mikati et al.[23]. Absolute burden for each respiratory hazardous air pollutant was calculated by census tract as the weighted average of the emissions over the block groups within each tract. Air pollutant emissions for the state of Louisiana were obtained from the 2017 National Emissions Inventory[24], and data for the census block groups and census tracts, including shape files and demographic characteristics, were obtained from the 2015-2019 American Community Survey[25]. Air pollutants included PM_2.5_ and hazardous air pollutants (HAPs) known to have respiratory health effects: 1,3-dichloropropene, 2,4-toluene-diisocyanate, acetaldehyde, acrolein, acrylic acid, arsenic, beryllium, cadmium, chlorine, chloroprene, chromium, diesel PM, formaldehyde, hexamethylene-1,6-diisocyanate, hydrazine, hydrochloric acid, naphthalene, nickel, polycyclic organic matter (POM), propylene, and triethylamine. Oil and gas wells and refineries, prevalent naphthalene sources, and a neoprene plant, a chloroprene source, fall within the hospital service area (Supplemental Figure B). We used the R software v4.0.5 for data organization (packages *dplyr, tidyr, bit65*, and *data.table*) and for the merger of geographic data with air pollution emissions data and output of shape files containing emissions burdens (packages *tigris, Hmisc, sp*, and *rgdal*). Emissions burdens were then assigned to 12,031 individual COVID-19 patients in the FMOLHS database for their census tract of residence. Bias minimization related to spatial assignment of emissions burdens is described in Mikati et al.[23].

Differences in population characteristics, including air pollutant burden, were first illustrated using summary statistics. Direct relationships of race with other demographic variables (age, sex, BMI, comorbidities, insurance status) or with disease-related variables (hospital admission, ICU admission, mortality) were tested via χ^2^ or ANOVA for categorical or continuous variables, respectively. Patient status was determined using hospital data for admission status, length of hospital stay, ICU status, and length of ICU stay. P-value < 0.05 for the χ^2^ or ANOVA test signified a significant difference between Black and White COVID-19 patients. Because comparisons were limited to these two racial groups, the final sample size was 11,331.

Associations of race with COVID-19 outcomes (hospital admissions, ICU admissions, mortality) were tested via a multistep process. First, a cross-table was created and χ^2^ testing was performed to analyze the relationship between race and health outcome. Mediation analysis was then used to test if a portion of the race-outcome relationship could be accounted for by an intermediate variable[26-28]. Potential mediators and covariates in the association between race and health effect were identified by testing for significant associations between each of the other variables with race and health effect, respectively. Associations of each variable with both race and health effect indicated that the variable is a potential mediator and so would be included in the mediation analysis. Variables associated with just health effect but not with race were identified as covariates and controlled in the mediation analysis. Significant mediators with the same sign as the total effect were considered as part of the racial differences explained by the mediator, while those with opposite sign suggested greater racial differences in effect considering the mediator. The R package *mma* was used to perform the mediation analysis[29].

We confirmed each of the criteria listed under for the STrengthening the Reporting of OBservational Studies in Epidemiology checklist for cross-sectional studies during completion of this manuscript.

## Results

Of the 11,331 patients, 5708 (50.4%) identified as non-Hispanic Black, and 5623 (49.6%) identified as non-Hispanic White (Table 1). In comparison, 33.8% of the population of Louisiana census tracts associated with patients’ residential addresses (referred to hereafter as the “patient population”) identified as Black, and 58.8% identified as non-Hispanic White. Census tract population data were available for 89% of patients. 6210 (54.8%) cases identified as Female, and 5119 (45.2%) identified as Male. On average, Black patients were 7.9 years younger than White patients. Black patients had a higher average BMI, but average BMI for both groups was in the obese range (BMI > 30). Length of hospital and ICU stays were both significantly higher among White patients, although that difference diminished for Medicare recipients and those without insurance. More Black patients had Medicaid or were uninsured, while more White patients had private insurance or Medicare. Among the twenty-two pollutants tested, emissions burden was significantly higher for Black patients in seventeen compounds and for White patients in three compounds, with no significant difference for two pollutants.

**Table 1.** Characteristics of the study population. When differences were statistically different, the characteristic and its highest value were shown in bold.

|  | Black | White |  |
| --- | --- | --- | --- |
| N | 5708 | 5623 |  |
|  | mean | mean | p<sup>+</sup> |
| **Age** | 48.5 | **56.4** | <2x10<sup>-16</sup> |
| **Hospital length of stay (d)** | 5.70 | **6.82** | 2.40x10<sup>-6</sup> |
| **ICU length of stay (d)** | 5.75 | **7.96** | 7.13x10<sup>-7</sup> |
| BMI | 31.7 | 30.6 | 0.128 |
| Hazardous Air Pollutants<sup>+</sup> |  |  |  |
| **Dichloro:1,3-dichloropropene** | 0.104 | **0.233** | 1.28x10<sup>-10</sup> |
| **2,4-toluene** | **0.0324** | 0.0143 | 9.10x10<sup>-8</sup> |
| **Acetaldehyde** | 4120. | **4640.** | 1.29x10<sup>-13</sup> |
| **Acrolein** | **495.** | 272. | <2x10<sup>-16</sup> |
| **Acrylic Acid** | **50.8** | 19.1 | 1.62x10<sup>-8</sup> |
| **Arsenic** | **2.33** | 1.00 | <2x10<sup>-16</sup> |
| **Beryllium** | **0.338** | 0.109 | <2x10<sup>-16</sup> |
| **Cadmium** | **3.63** | 0.842 | <2x10<sup>-16</sup> |
| **Chlorine** | **3680** | 216. | <2x10<sup>-16</sup> |
| **Chloroprene** | **231.** | 50.9 | <2x10<sup>-16</sup> |
| **Chromium** | **2.46** | 0.443 | <2x10<sup>-16</sup> |
| **Diesel PM** | **0.293** | 0.0481 | <2x10<sup>-16</sup> |
| **Formaldehyde** | **4430.** | 1830. | <2x10<sup>-16</sup> |
| **HCl** | **21700** | 2720. | <2x10<sup>-16</sup> |
| **Hexamethylene 6-diisocyanate** | 0.410 | **2.89** | <2x10<sup>-16</sup> |
| Hydrazine | 0.00231 | 0.0152 | 0.565 |
| **Naphthalene** | **2280.** | 255. | <2x10<sup>-16</sup> |
| **Nickel** | **95.3** | 23.1 | <2x10<sup>-16</sup> |
| **PM2.5** | **83.0** | 16.1 | <2x10<sup>-16</sup> |
| **Polycyclic organic matter** | **0.0231** | 0.00793 | 1.47x10<sup>-5</sup> |
| Propylene | 8.90 | 6.44 | 0.289 |
| **Triethylamine** | **46.6** | 19.0 | <2x10<sup>-16</sup> |
| <sup>+</sup> p-values were calculated using square root transformed data to normalize the data distribution |  |  |  |

For the study duration, hospital admissions were significantly higher among White patients, while ICU admissions were significantly higher among Black patients. Compared to their share of the patient population, Black patients were overrepresented among hospital admissions by 28%, among ICU admissions by 43%, and among total COVID-19 patients by 38% (Table 2). Hospital and ICU admissions significantly exceeded the share of the population for Black patients by 86% and 89%, respectively during the first wave and by 40% and 56%, respectively during the second wave. By the third wave, the proportions of hospital and ICU admissions were higher among White patients with a significant χ^2^, but the proportion of hospital and ICU admissions among Black patients were 16% and 36% greater than the share of the population identifying as Black.

**Table 2.** Count tables for χ^2^ analysis for each wave of the study and for the full study period. Equitable Black and equitable White indicate the ratio of the share of the population of patients in each group compared to the number of patients that should be in each group based on the proportion of each group in Louisiana census tracts sending patients to the FMOLHS.

|  | HA | ICU | No HA | Death | No Death |
| --- | --- | --- | --- | --- | --- |
| First Wave: March 1, 2020-June 10, 2020 |  |  |  |  |  |
| Black | **256** | **192** | **314** | **107** | **655** |
| Equitable Black | **1.86** | **1.89** | **2.10** | **1.78** | **1.99** |
| White | 121 | 86 | 96 | 58 | 245 |
| Equitable White | 0.51 | 0.49 | 0.37 | 0.55 | 0.43 |
| p | 0.0149 |  |  | 0.0475 |  |
| Second Wave: June 11, 2020-October 6, 2020 |  |  |  |  |  |
| Black | **307** | **188** | **645** | 58 | **1082** |
| Equitable Black | **1.40** | **1.56** | **1.64** | **1.28** | **1.57** |
| White | 294 | 143 | 433 | 66 | 804 |
| Equitable White | 0.77 | 0.68 | 0.63 | 0.84 | 0.67 |
| p | 2.43x10<sup>-3</sup> |  |  | 0.0269 |  |
| Third Wave: October 7, 2020-June 30, 2021 |  |  |  |  |  |
| Black | 619 | 287 | **1178** | 83 | 1921 |
| Equitable Black | **1.16** | **1.36** | **1.42** | 0.98 | **1.34** |
| White | **843** | **290** | 1089 | **148** | **2015** |
| Equitable White | 0.91 | 0.79 | 0.76 | **1.01** | 0.81 |
| p | 5.47x10<sup>-8</sup> |  |  | 1.85x10<sup>-4</sup> |  |
| Fourth Wave: July 1, 2021-August 31, 2021 |  |  |  |  |  |
| Black | 439 | 59 | 1224 | 41 | 1590 |
| Equitable Black | **1.16** | 0.81 | **1.24** | 0.99 | **1.18** |
| White | **601** | **141** | **1486** | 72 | 2087 |
| Equitable White | 0.91 | **1.11** | 0.86 | 1.00 | 0.89 |
| p | 5.31x10<sup>-5</sup> |  |  | 0.169 |  |
| Full Study Period: March 1, 2020-August 31, 2021 |  |  |  |  |  |
| Black | 1621 | **726** | **3361** | 289 | **5248** |
| Equitable Black | **1.28** | **1.43** | **1.42** | **1.25** | **1.38** |
| White | **1859** | 660 | 3104 | **344** | 5151 |
| Equitable White | 0.84 | 0.75 | 0.76 | 0.86 | 0.78 |
| p | 5.04x10<sup>-7</sup> |  |  | 0.0209 |  |

Information regarding mortality (patients who expired while at the hospital or within 7 days of discharge) was available for 11,032 (97.3%) cases (Table 2). For the study duration, the proportion of those who died was significantly higher for White patients, based on the χ^2^ test, but the proportion of Black patients who died was still 25% greater than the proportion of Black people in the Louisiana census tracts sending patients to FMOLHS. The proportion of patients who died was nearly 65% for Black patients during the first wave, with the share of the patient population that is Black overrepresented by 78% but was significantly higher for White patients during the second and third waves and not significantly different in the fourth wave. During the second wave, mortality among Black patients was still 28% higher than the share of patient population identifying as Black.

Age and, with smaller contribution, comorbidities were significant mediators of the race-hospitalization relationship (Figure 1) for the entire study period. Age and comorbidities were also consistently significant mediators for each wave. Naphthalene and arsenic were significant mediators of the race-hospitalization relationship for the duration of the study. Naphthalene was not a significant mediator for any of the waves, and arsenic was only for the fourth wave. PM_2.5_ and chromium added uncertainty to the race-hospital admissions relationship, because the different sign of these mediation coefficients widened the confidence intervals around the total effect. Cadmium added uncertainty to the race-hospitalization relationship for the third wave. Several studies[7,11-13] found associations of PM_2.5_ with COVID-19 using data from the first few months of the pandemic, but they either used a nationwide domain or studied different parts of the country. Terrell and James[15] calculated a correlation of 0.21 for PM_2.5_ concentration with COVID-19 mortality for Louisiana, and Xu et al.[30] noted for a study of COVID-19 in Texas that PM_2.5_ concentrations were not associated with COVID-19 mortality.

**Figure 1:**
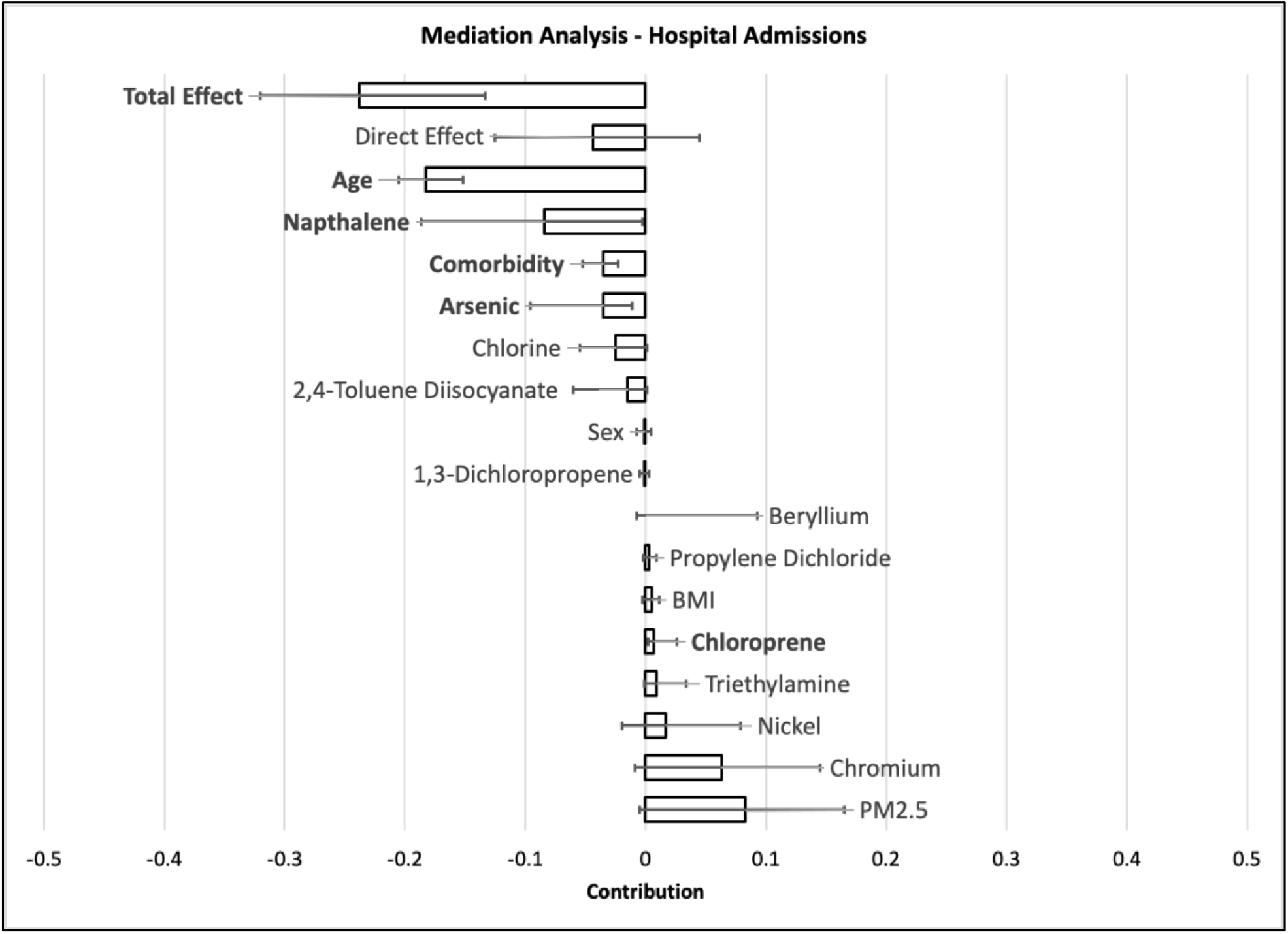
Mediation analysis results for hospital admissions. Whiskers indicate the 95% confidence interval around the mediation effect, with each tested mediator shown by a column. Statistically significant effects are bolded.

The model for race-ICU admission for the entire study period (Figure 2) included a direct effect that was larger than and opposite in sign to total effect, widening the confidence interval around total effect to suggest uncertainty. This model also contained age as a mediator and, with smaller magnitude, comorbidity and sex. Chloroprene and naphthalene emissions were both significant mediators, while PM_2.5_ and chromium emissions appeared to widen the confidence intervals around total effect. Age was a mediator of the race-ICU admission effect during each wave. During the third wave, the total effect between race and ICU admission was near zero, but there was a positive direct effect and positive indirect effect of PM_2.5_ emissions balanced by negative indirect effects of age, cadmium emissions, and nickel emissions. The fourth wave produced a large total effect for the race-ICU admission model that included a direct effect comprising more than half of the total effect and indirect effects from age, insurance status, sex, and emissions of POM.

**Figure 2:**
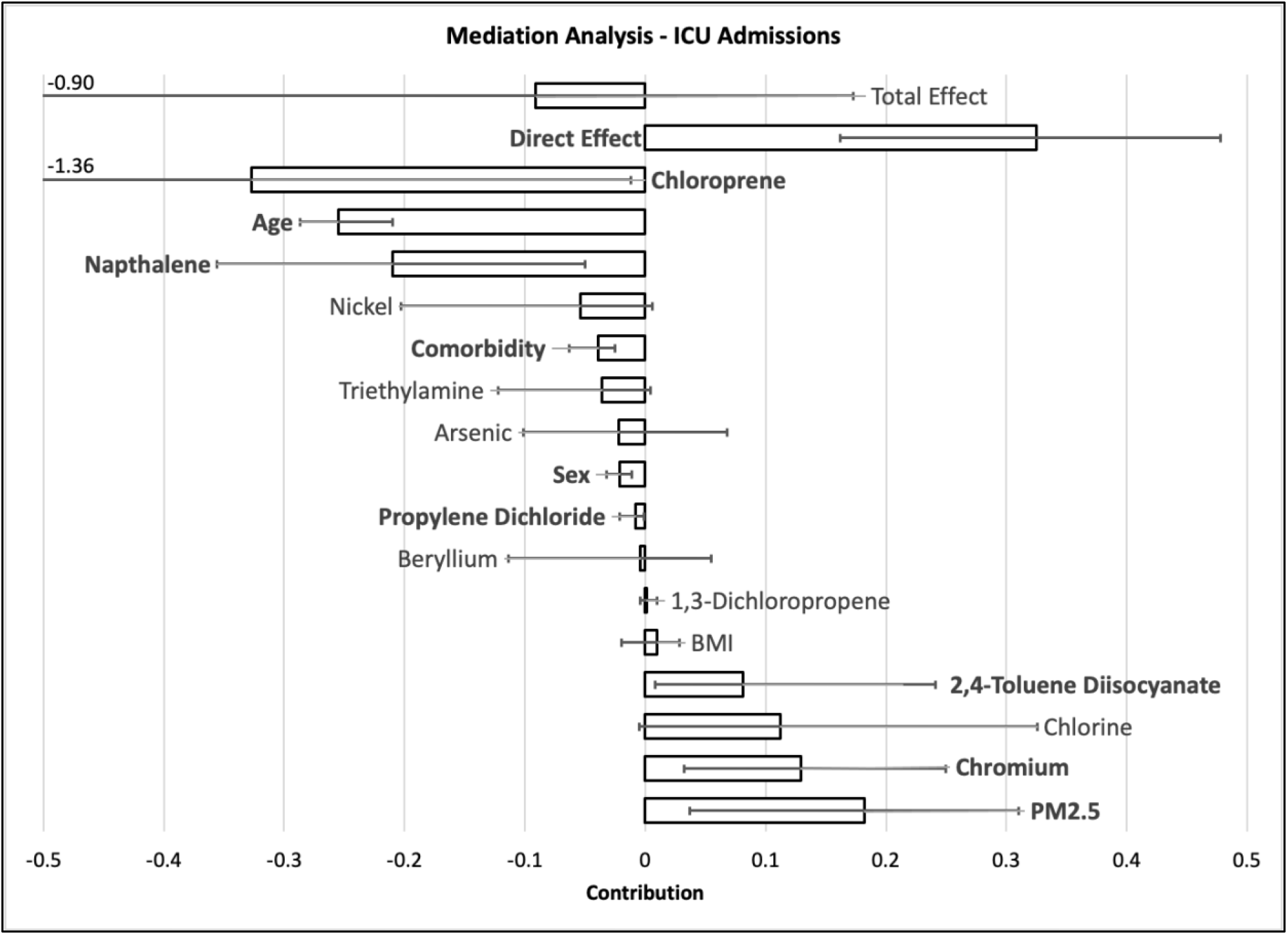
Mediation analysis results for ICU admissions. Whiskers indicate the 95% confidence interval around the mediation effect, with each tested mediator shown by a column. Where the lower confidence interval goes beyond the data range shown on the page, the lower bound is provided numerically on the graph. Statistically significant effects are bolded.

Mediation analysis results indicate that, for the total duration and each wave, age was consistently a significant mediator of the race-mortality relationship (Figure 3), consistent with Cronin and Evans[31]’s finding of higher COVID-19 mortality for Black males and females for every age group (0-44 y, 45-64 y, 65-74 y, 75+ y) with a greater effect of age than race or sex. Sex and comorbidities had smaller indirect effects for the entire study period but were still significant. Naphthalene was identified as a mediator of the race-mortality relationship for the total duration, while hydrochloric acid added uncertainty to the assessment of mediation. Naphthalene was identified as a potential mediator during the first wave but was not significant and added uncertainty to that model. POM was a significant mediator of the race-mortality relationship during the fourth wave. POM emerged as a potential mediator in the total duration model but was of small magnitude.

**Figure 3:**
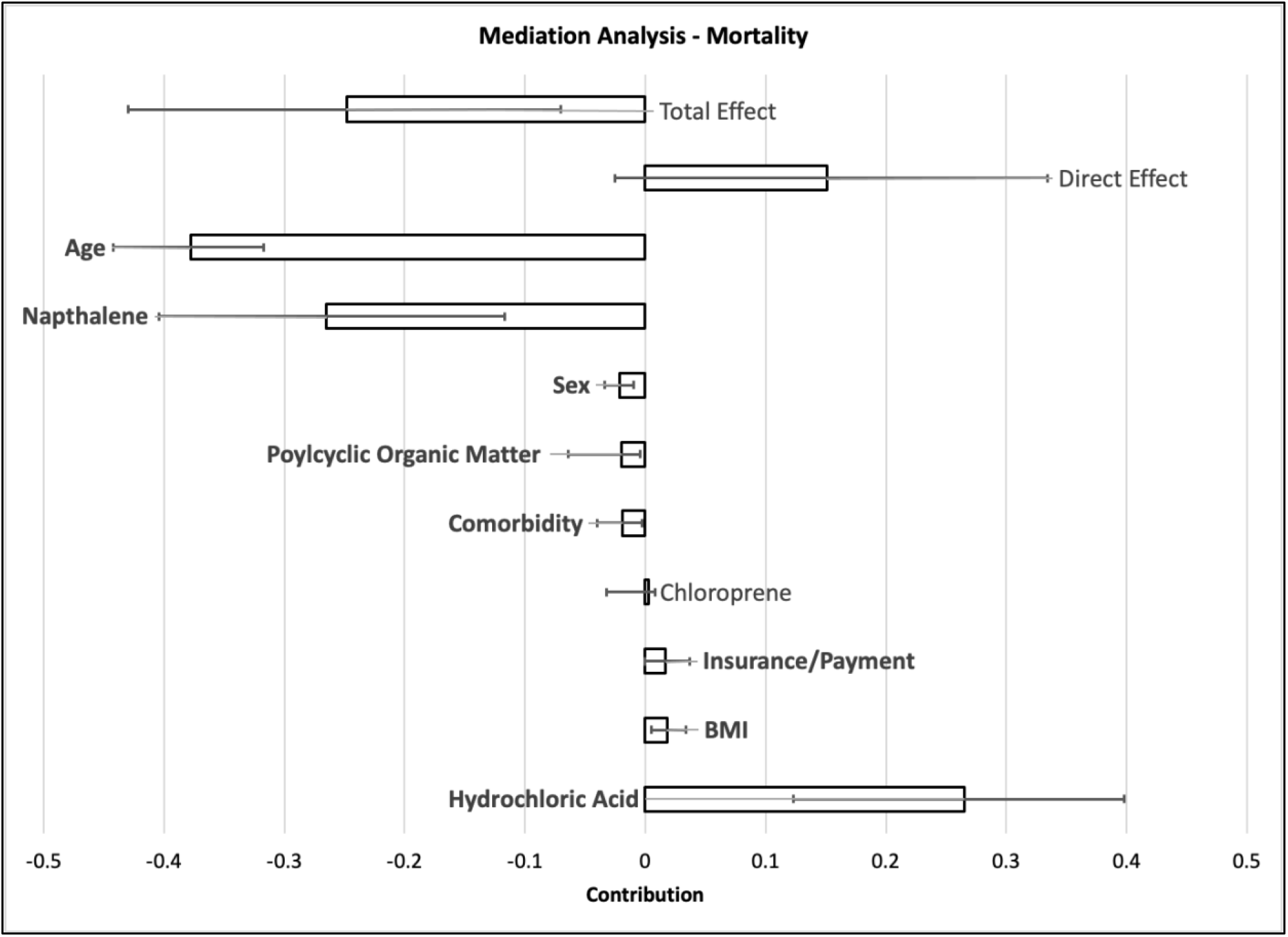
Mediation analysis results for mortality. Whiskers indicate the 95% confidence interval around the mediation effect, with each tested mediator shown by a column. Statistically significant effects are bolded.

There were some limitations specific to this dataset. We used data from one hospital system. This selective population was not representative of all Louisiana COVID-19 hospitalizations and could have imposed selection bias. The most recent HAP emission data were from 2017. Additionally, vaccination status was not included in the dataset but could have affected severe outcomes during the last two waves.

Mediation analysis showed a clear relationship between race and outcome at the beginning of the pandemic, but race appeared less influential over time. Mediation analyses highlighted the uncertainty in the race-outcome relationships across waves. Although several air pollutants were associated with race, with higher emissions burdens among predominantly Black census tracts, air pollution did not appear to consistently mediate the race-outcome relationship for most waves. Uncertainties in the mediation analyses raise questions about unmeasured confounding. VanderWeele[32] asserted four necessary assumptions for mediation analysis: 1) control for confounding of the exposure-outcome relationship, 2) control for confounding of the mediator-outcome relationship, 3) control for confounding of the exposure-mediator relationship, and 4) no confounder of the mediator-outcome relationship is affected by the exposure. The first three were accomplished through the process of checking for significant associations among the exposure, potential mediator, and outcome. However, the final assumption is more difficult to enforce for this study given that long-standing racialization may introduce other, uncontrolled factors[33]. Similarly, it is difficult to ascertain whether any mediators were omitted from the analysis. Additionally, exposure measurement error or exposure misclassification has the potential to weaken the associations between the exposure and mediators. In the case of the HAP burdens, Mikati et al.[23] sought to control this by testing different assignment radii and found little difference. Use of census tract-level assignments also helps to localize the exposure estimates.

## Conclusions

A complicated picture of relationships between race and COVID-19 hospitalizations, ICU admissions, and mortality emerges from these results. For the entire study period, hospitalization and mortality rates among those diagnosed with COVID-19 were greater for White patients than for Black patients, while ICU admission rates were higher for Black patients. These proportions shifted towards White patients and were significant by late 2020. But, the proportion of those diagnosed with COVID-19 as well as those hospitalized, admitted to the ICU, and died remained disproportionately high for Black patients compared with the patients’ residential areas, despite the 10-year age difference between Black and White patients.

Among the population of hospitalized COVID-19 patients, most of the effect of race could be explained by mediators. Age was the strongest mediator, accounting for the largest share of the indirect effect. In each wave, the average age of Black patients was 8-9 years younger than the average age of White patients. In fact, life expectancy for Black Louisiana residents is 3.4 years shorter than for White Louisiana residents[34]. These factors make it difficult to disentangle the effect of race from the effect of age.

Findings that naphthalene and chloroprene acted as mediators of race for ICU admissions and that naphthalene acted as a mediator for hospitalizations and mortality were not surprising because their burdens among Black patients were 8.9 and 4.5 times higher, respectively than for White patients. Our results imply that policies to improve environmental conditions – especially among Louisiana’s predominantly Black communities – may have lessened inequities in COVID-19 impacts.

## Data Availability

Data are not publicly available.

