## Supplemental Material for "Risk factors among Black and White COVID-19 patients from a Louisiana Hospital System, March, 2020 – August, 2021"

**Contents**

**FIGURES**……………………………………………………………………………………3
**Supplemental Figure A:** Hypothetical causal pathways showing that the association between race and COVID-19 may be mediated by comorbidities, insurance status, and pollution exposure……………………………………………………………………………………... 3
**Supplemental Figure B:** Locations of a chloroprene point source and of disperse naphthalene sources in the region of Southern Louisiana feeding patients to the Franciscan Missionaries of Our Lady Health System...…………………………………………………………………….. 4

**FIGURES**

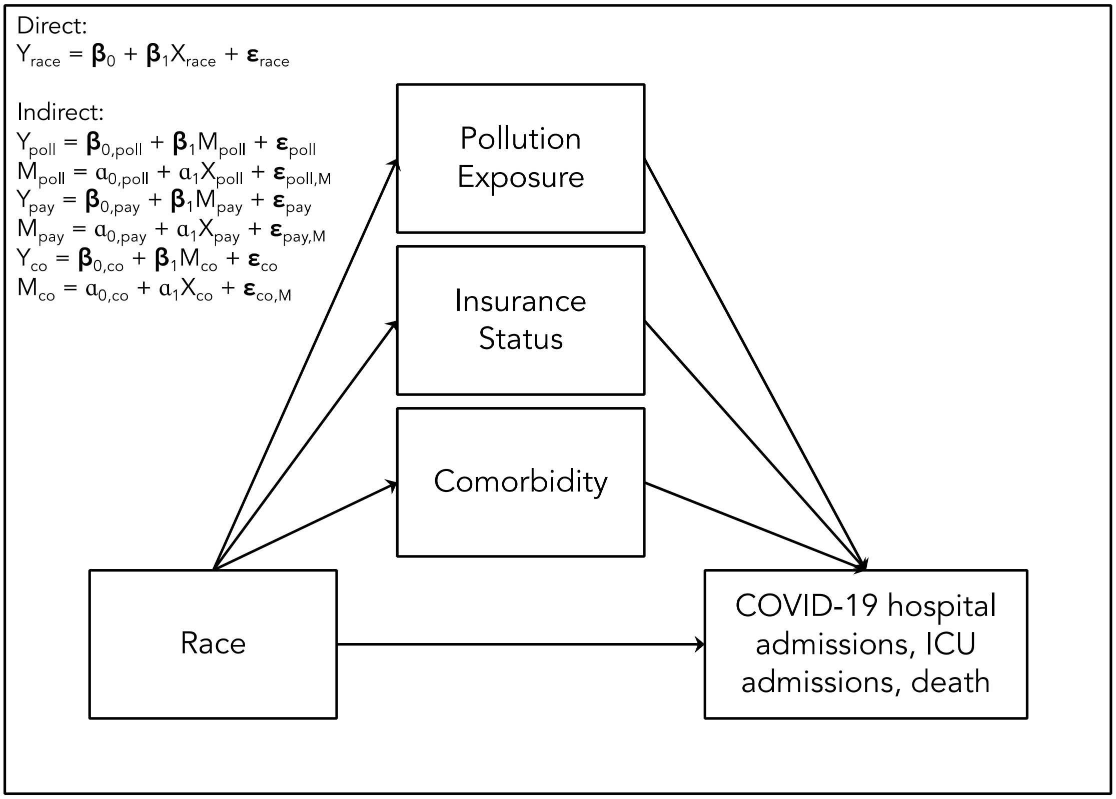


**Supplemental Figure A:** Hypothetical causal pathways showing that the association between race and COVID-19 may be mediated by comorbidities, insurance status, and pollution exposure.

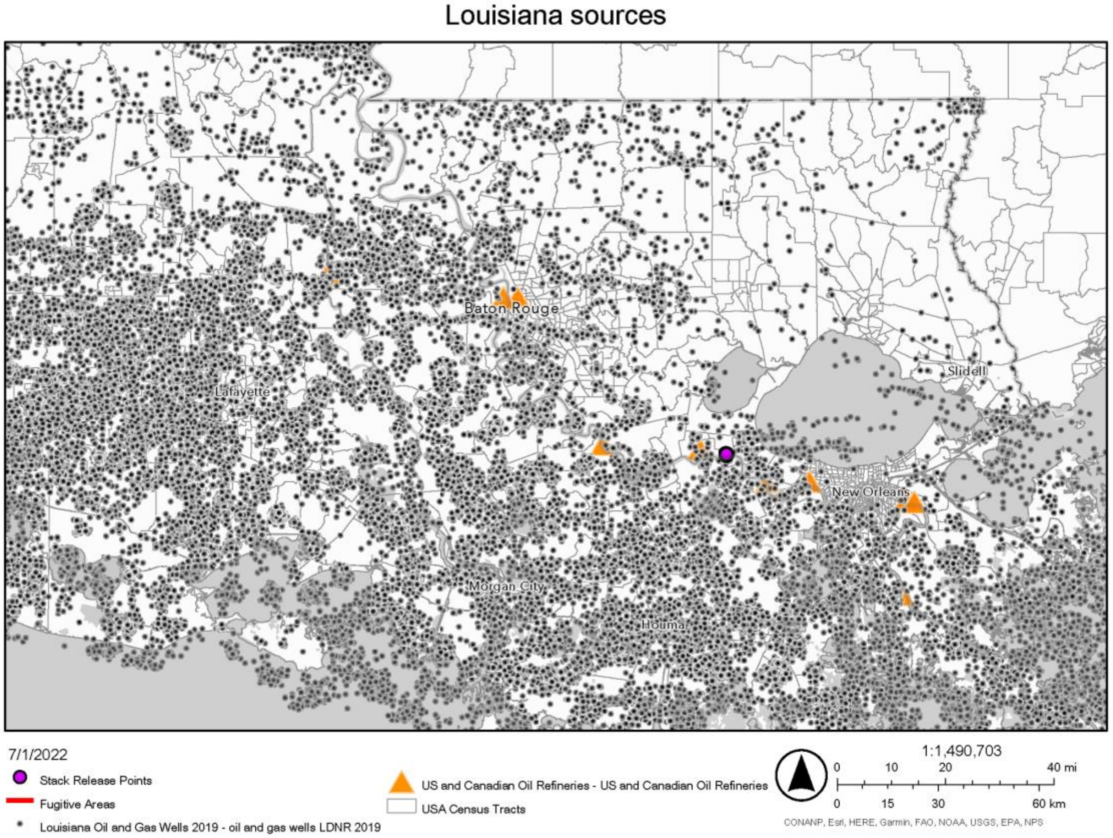


**Supplemental Figure B:** Locations of a chloroprene point source (purple stack release from one major emitting facility) and of disperse naphthalene sources (black dots to show oil and gas wells, orange triangles to show oil refineries) in the region of Southern Louisiana feeding patients to the Franciscan Missionaries of Our Lady Health System.
